# Safety and immunogenicity of a heterologous booster of protein subunit vaccine MVC-COV1901 after two doses of adenoviral vector vaccine AZD1222

**DOI:** 10.1101/2021.12.10.21267574

**Authors:** Shu-Hsing Cheng, Yi-Chun Lin, Cheng-Pin Chen, Chien-Yu Cheng

## Abstract

We report the interim safety and immunogenicity results in participants administrated with a booster dose of protein subunit vaccine MVC-COV1901 at 12 or 24 weeks after two doses of AZD1222 (ChAdOx1 nCoV-19). In subjects fully vaccinated with two doses of AZD1222, waning antibody immunity was apparent within six months of the second dose of AZD1222. At one month after the MVC-COV1901 booster dose, anti-SARS-CoV-2 spike IgG antibody titers and neutralizing antibody titers were 14- and 8.6-fold increased, respectively, when compared to the titer levels on the day of the booster dose. We also observed 5.2- and 5.6-fold increases in neutralizing titer levels against wildtype and Omicron variant pseudovirus after the booster dose, respectively. These interim results support the use of MVC-COV1901 as a heterologous booster for individuals vaccinated with AZD1222.

## Introduction

The COVID-19 pandemic is claiming millions of lives worldwide and the negative economic and public health impacts may have been far greater without the currently approved vaccines including the two mRNA- and two adenovirus-based anti-SARS-CoV-2 vaccines from Moderna or Pfizer/BioNTech and Oxford/AstraZeneca or Johnson & Johnson, respectively. These vaccines are highly efficacious in preventing serious disease, hospitalization and death and have demonstrated good safety records overall [1]. Nevertheless, as is the case with all vaccines, there are rare but severe safety signals that have led to limited applicability to specific patient subgroups. Examples are the mRNA vaccines that have been associated with rare events of myocarditis in younger males and thrombosis associated with adenoviral vector vaccines [2, 3].

New evidence shows challenges that comes with an increased infectiousness and immune evasion of emerging variants of concern (VoCs) including Delta and Omicron variants, and the discovery of a relatively short duration of protection conferred by COVID-19 vaccines [4, 5]. These are concerns that developed with the resurgence of cases and the observation that vaccinated people show increasing rates of infection starting about 6 months post-vaccination [6]. Thus, consensus is building that long term COVID-19 control may be achieved by booster shots that may well become annual events [7].

AstraZeneca AZD1222 has the lowest cost amongst approved vaccines and because there is no need for an extreme-cold chain infrastructure such as the one needed for the mRNA vaccines, it is ideal for use in lower and middle-income countries [8, 9]. However, AZD1222 has been found to be of weaker immunogenicity compared to other widely used mRNA vaccines, but boosters with different vaccines (heterologous boosting) may compensate for this potential deficiency as shown in the COV-BOOST trial in the UK [10]. If proven, this may lead to a truly cost-effective vaccine-booster combination.

MVC-COV1901 is a subunit vaccine based on the stable prefusion spike protein (S-2P) of SARS-Cov-2 adjuvanted with CpG 1018 and aluminum hydroxide and has been approved for use after a large phase 2 clinical trial demonstrated favorable safety and immunogenicity profiles [11]. An EUA was granted to MVC-COV1901 since July, 2021 and the vaccine was rolled-out in Taiwan since the end of August, 2021. Based on a post-marketing safety surveillance system run by the Taiwanese Centers for Disease Control, no alarming safety signals have been reported for MVC-COV1901 [12]. Thus, we are investigating if MVC-COV1901 boosters can attain optimal immunogenicity after waning immunity from initial immunization(s).

In this study we report results from an MVC-COV1901 booster shot after two initial immunizations with AZD1222. We used data to quantify anti-SARS-CoV-2 spike IgG antibody titers and neutralizing antibody titers boosted by an MVC-COV1901 booster dose and describe the initial safety findings of the booster dose. We also investigated the enhancement in immunogenicity of booster dose of MVC-COV1901 against the Omicron variant pseudovirus.

## Methods

### Study design

This study is a parallel, prospective, randomized, open-label, clinical study to evaluate the immunogenicity, safety, and tolerability of MVC-COV1901 as a booster vaccine in participants that have received two doses of AZD1222. Two hundred and one healthy adults from age of 23 to 66 years that have received two doses of AZD1222 within 6 months of study initiation were randomized into two groups. Participants in Group A were scheduled to receive a booster dose of MVC-COV1901 (15 mcg of S2-P adjuvanted with 750 mcg of CpG 1018 and 375 mcg of aluminum hydroxide) administered intramuscularly 12 weeks after the last dose of AZD1222, whereas those in Group B were scheduled to receive a booster dose of MVC-COV1901 administered intramuscularly 24 weeks after the last dose of AZD1222. For analysis of the immunogenicity, 73 respondents were selected. The study design and flowchart is presented in Figure 1. The timeline of the study is as outlined in Figure 2. Day 1 is defined as the day that Group A participants received the dose of MVC-COV1901 booster. Therefore, in Group B participants, the MVC-COV1901 is administered on Day 85. Blood samples were taken at immunization and during additional study visits (Days 29, 85 and 169).

**Figure 1.**
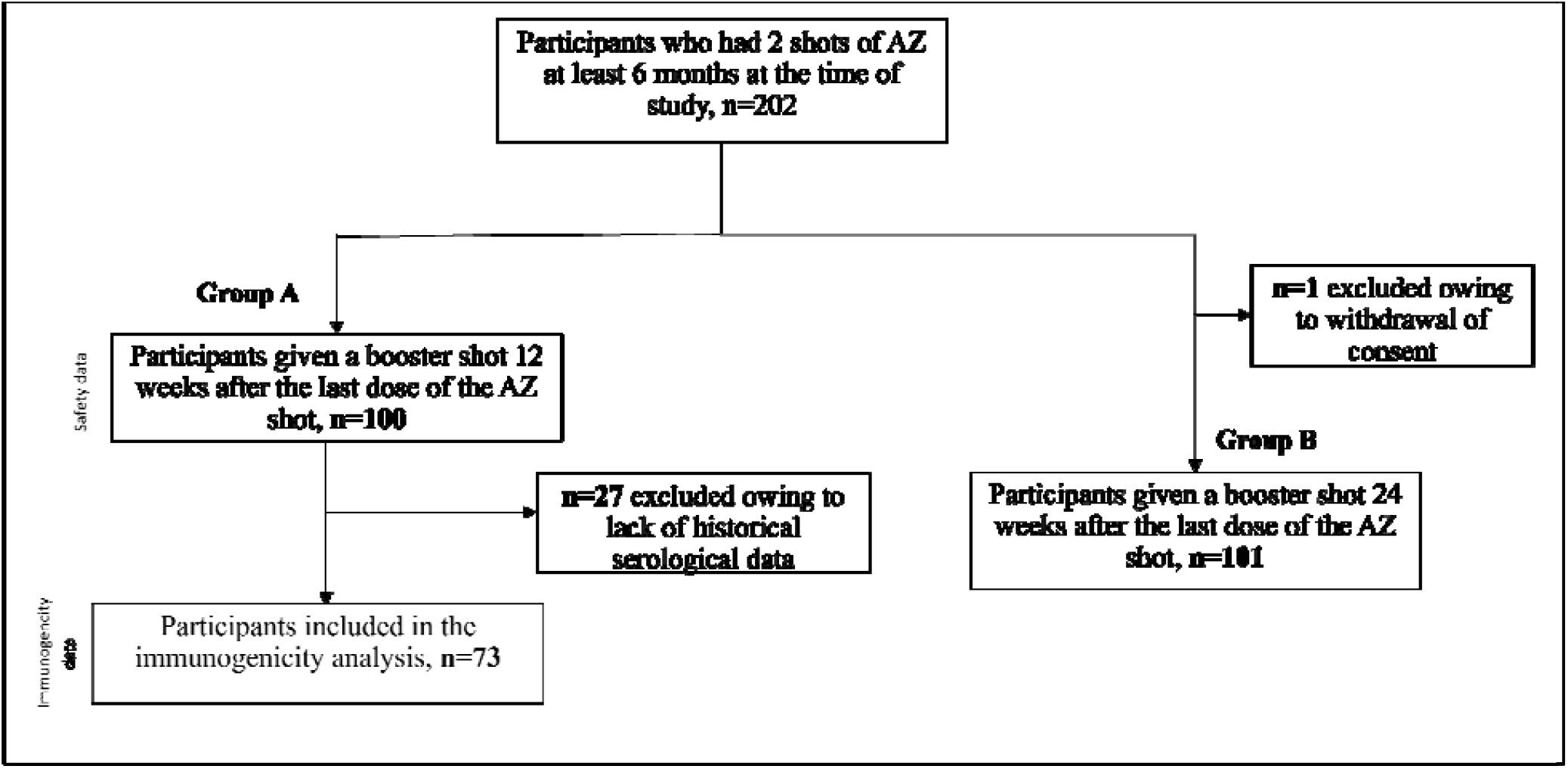
Study design and flowchart

**Figure 2.**
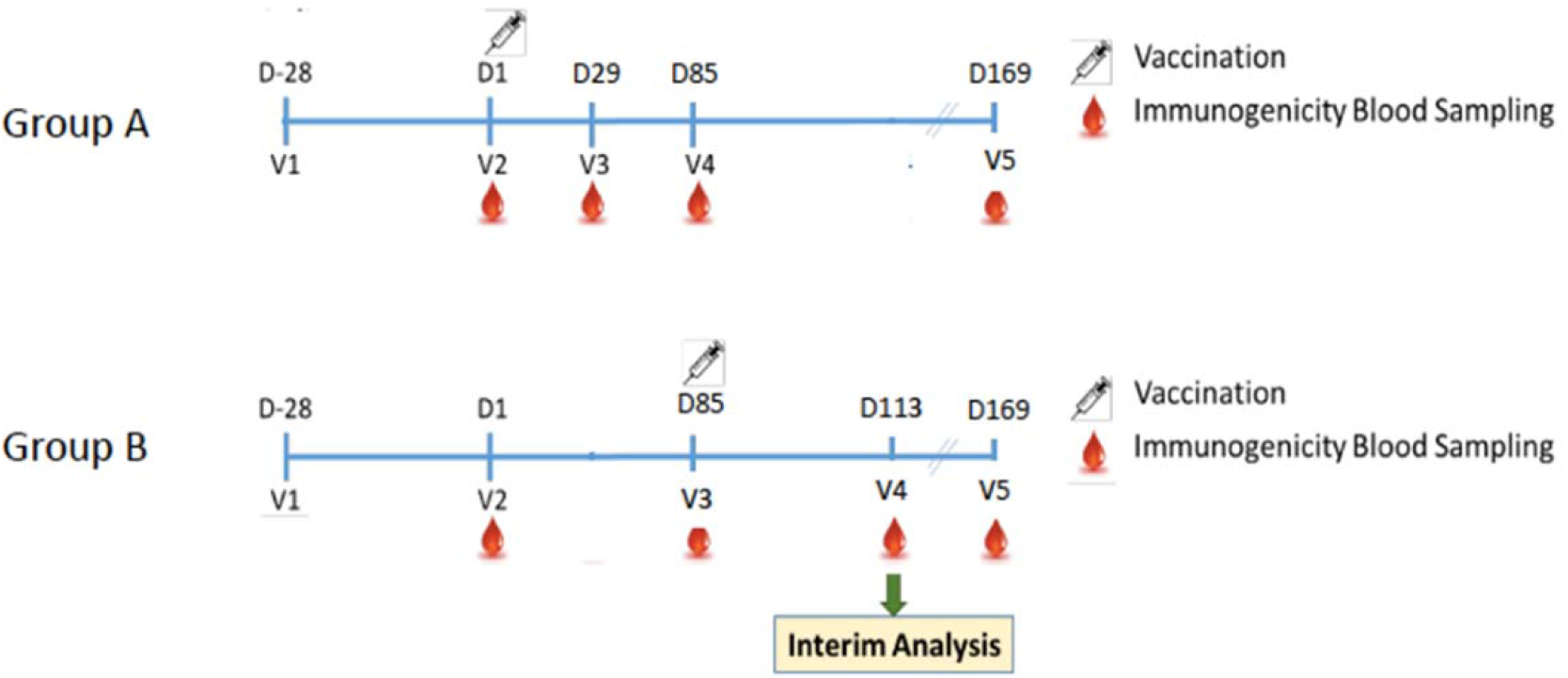
Study timeline and key time points.

### Outcomes

Safety was assessed by monitoring for solicited and unsolicited adverse events (AEs) for the first 14 days after the booster dose, and 28 days and 12 weeks after the booster dose. Immunogenicity was assessed by measuring anti-SARS-CoV-2 spike IgG antibody and live-SARS-CoV-2 neutralizing assay as previously performed [11]. An eCRF was used to record the actual date and time of sample collection. Unique sample identification was used to always maintain blinding at the laboratory and to allow for automated sample tracking and storage.

### Laboratory methods

The detection and characterization of antigen-specific immunoglobulin was performed by a central laboratory using a validated enzyme-linked immunosorbent assay ELISA) method using customized 96-well plates coated with S-2P antigen.

Live-SARS-CoV-2 neutralization assay was performed as previously with wildtype SARS-CoV-2, Taiwan CDC strain number 4 (hCoV-19/ Taiwan/4/2020; GISAID accession ID EPI_ISL_411927) [11]. The serum samples underwent a total of eight two-fold dilutions, starting from a 1:8 dilution to a final dilution of 1:1024. Diluted serum samples were then mixed with an equal volume of 100 TCID_50_ per 50 μL of virus and incubated at 37°C for 1 hour. After incubation, the mixture was added Vero E6 cells and incubated at 37°C in a 5% CO2 incubator for 4–5 days. The neutralising titer (NT_50_) was estimated as the reciprocal of the highest dilution capable of inhibiting 50% of the cytopathic effect. The NT_50_ results were calculated with the Reed-Muench method. Anti-spike IgG titers and neutralizing antibody titers were converted to the WHO Standardized Unit, BAU/mL and IU/mL, respectively. The conversion is based on the WHO validated NIBSC reference panel. Results are expressed as geometric mean titer (GMT) and converted to binding antibody units (BAU/mL) for IgG titer and international units (IU/mL) for neutralizing antibody titer as we have performed in our phase 2 clinical study [11].

Pseudovirus with spike proteins of wildtype and Omicron variant were constructed and neutralization assays performed. Two-fold serial dilution of serum samples were mixed with equal volume of pseudovirus and incubated at 37°C for 1 hour before adding to the HEK-293-hAce2 cells. Fifty percent inhibition dilution titers (ID50) were calculated with uninfected cells as 100% neutralization and cells transduced with virus as 0% neutralization. The mutations for Omicron variant used in the spike sequence for pseudovirus construction are: A67V, del69-70, T95I, G142D, del143-145, del211, L212I, ins214EPE, G339D, S371L, S373P, S375F, S477N, T478K, E484A. Q493R, G496S, Q498R, N501Y, Y505H, T547K, D614G, H655Y, N679K, P681H, D796Y, N856K, Q954H, N969K, L981F.

## Results

In this interim report, we describe the findings from the Group A as Group B is still in progress at the time of writing. Table 1 summarizes the demographic characteristics of the sample. The median age of Group A is 40 years with an Interquartile range (IQR) of 13.0 years, while approximately 32% are males. In terms of Body Mass Index (BMI) approximately 13% in the group have BMI greater than 30 kg/m^2^. Median BMI is 23.97 kg/m^2^ with an IQR of 6.03. Figures 3 and 4 involved analyzing data from a subsample of 73 respondents.

**Table 1.** Demographic profile of respondents

| Item\MVC lot | Vaccine group |  | p-value |
| --- | --- | --- | --- |
|  | Group A | Group B |  |
| □ Age (years) |  |  |  |
| N (Missing) | 100 (0) | 101 (0) | 0.673 |
| Mean (SD) | 41.4(10.3) | 42.03 (10.9) |  |
| Median (IQR) | 40 (13.0) | 40 (16.0) |  |
| Q1~Q3 | 34.5~47.5 | 35~51 |  |
| Min~Max | 24~66 | 23~66 |  |
| **Gender** |  |  |  |
| N (Missing) | 100 (0) | 101 (0) |  |
| Male | 32 (32.0) | 29 (28.7) | 0.612 |
| Female | 68 (68.0) | 72 (71.3) |  |
| **BMI (kg/m<sup>2</sup>)** |  |  |  |
| N (Missing) | 100(0) | 101(0) |  |
| Mean (SD) | 24.4(4.4) | 24.6 (4.2) |  |
| Median (IQR) | 23.97(6.03) | 23.8 (6.3) | 0.74 |
| Q1~Q3 | 21.2~27.2 | 21.3~27.6 |  |
| Min~Max | 17.6~35.3 | 17.99~37.8 |  |
| **BMI group** |  |  |  |
| N (Missing) | 100 (0) | 101 (0) |  |
| <30 kg/m<sup>2</sup> | 87 (87.0) | 88 (87.1) | 0.978 |
| >=30 kg/m<sup>2</sup> | 13 (13.0) | 13 (12.9) |  |
| **Comorbidity Category** |  |  |  |
| N (Missing) | 100 (0) | 101 (0) |  |
| Yes | 32 (32.0) | 25 (24.8) | 0.254 |
| No | 68 (68.0) | 76 (75.3) |  |

(1) Abbreviations: N=number of subjects in PPI population; SD=standard deviation; Q1=first quartile (25th percentile); Q3=third quartile (75th percentile); IQR=interquartile range; BMI=Body Mass Index; HIV=Human Immunodeficiency Virus.

**Figure 3.**
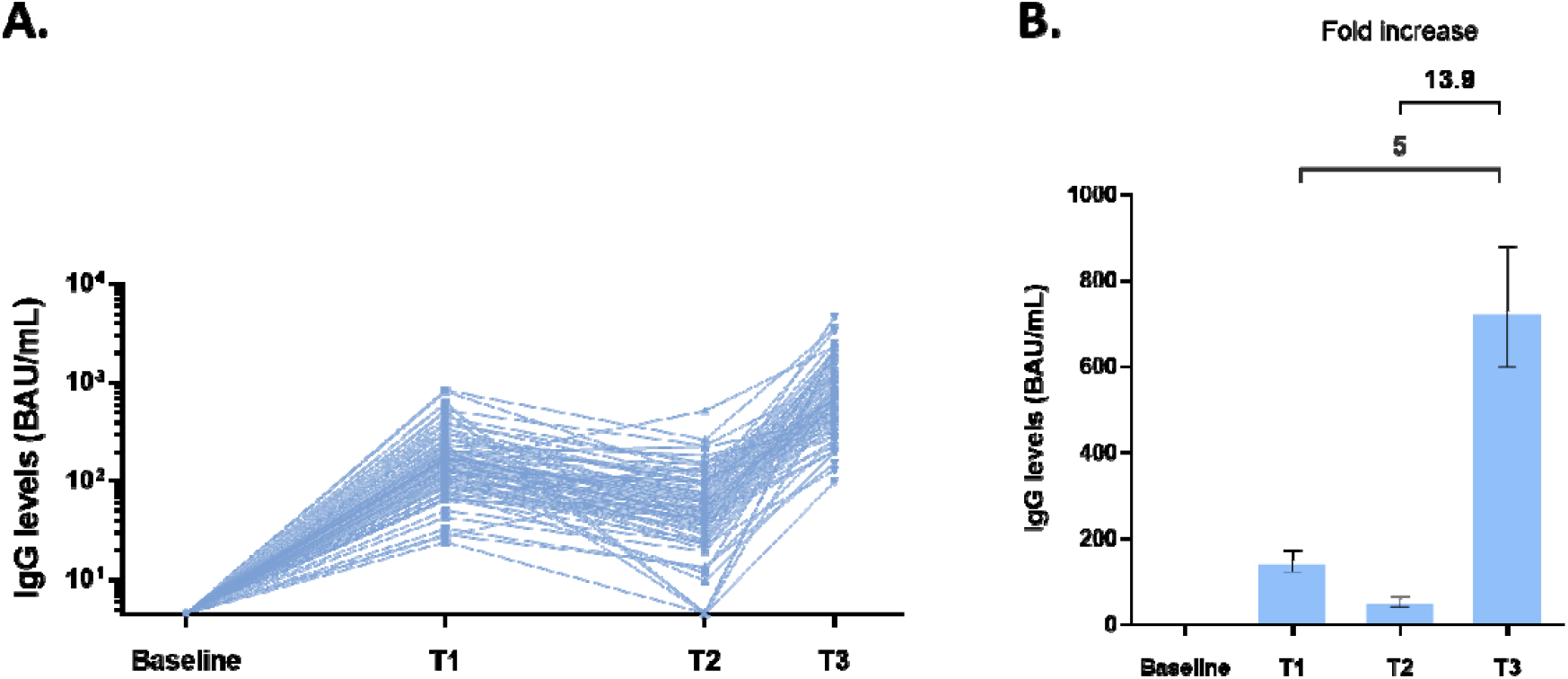
Anti-SARS-CoV-2 spike IgG titers at different time points. Results were presented by A) blue line/dot for each individual IgG level and B) bars representing geometric mean IgG titer with error bars for 95% confidence interval values. T1: 28 days after second dose of AZD1222; T2: On the day of MVC-COV1901 booster dose; T3: 28 days after MVC-COV1901 booster dose

**Figure 4.**
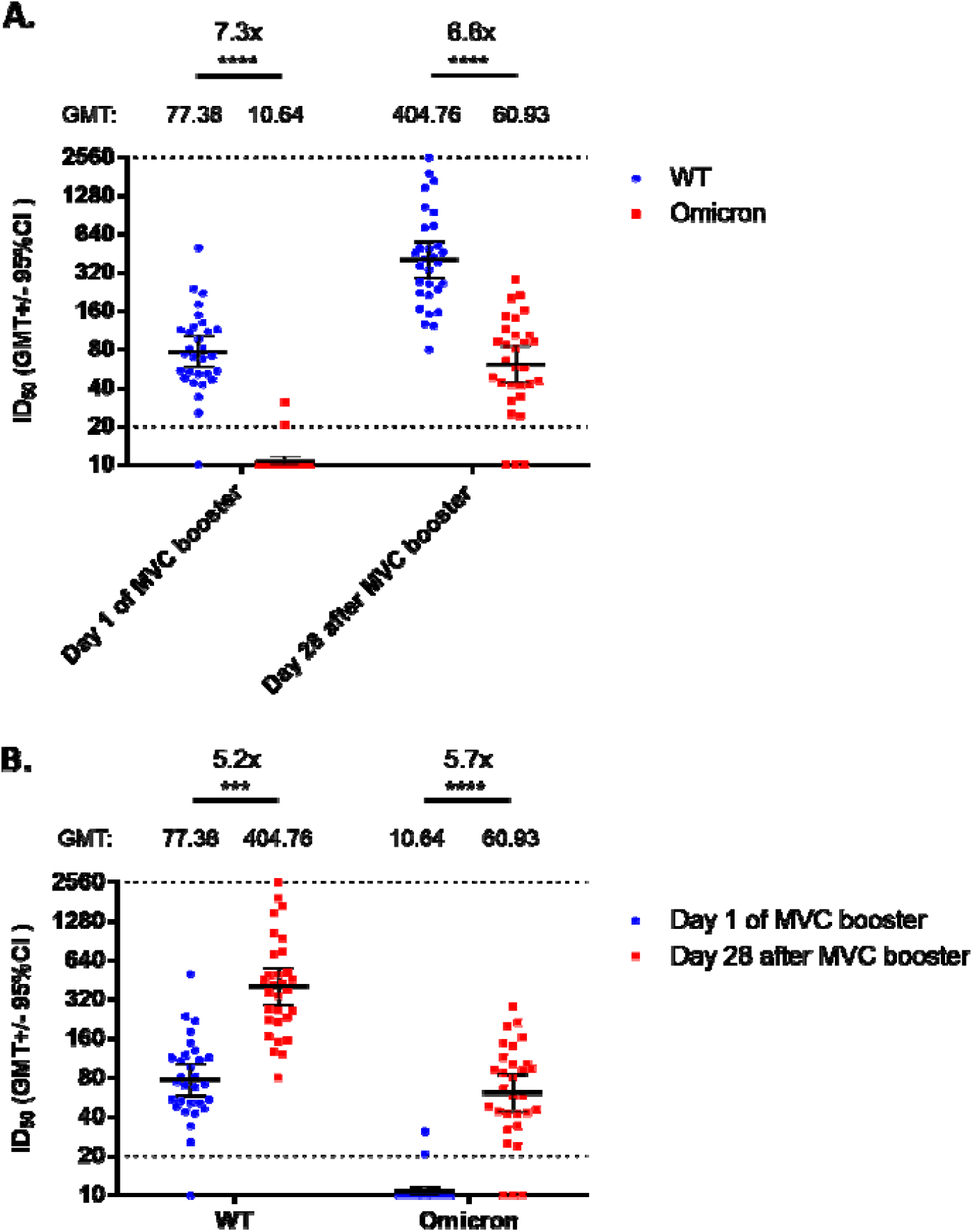
Neutralizing assay against wildtype and Omicron variant pseudoviruses. Serum samples on the day of MVC booster dose and day 28 after MVC booster dose from 30 randomly selected individuals of the immunogenicity analysis subset were taken. A) grouped according to the time of sampling; B) grouped according to pseudovirus type. Results were presented by horizontal bars representing geometric mean titer with error bars for 95% confidence interval values. Statistical significance was calculated with Kruskal-Wallis test with corrected Dunn’s multiple comparisons test. * = p < 0.05, ** = p < 0.01, *** = p < 0.001, **** = p < 0.0001

The results of immunogenicity are summarized in Table 1 and Figure 3. Figure 3 shows the anti-spike IgG titers at T1: 28 days after the second AZD1222 shot; T2: day of vaccination for the MVC-COV1901 booster shot within three months after the last AZD1222 dose; and T3: one month after a booster dose of MVC-COV1901. At T1, the anti-spike IgG geometric mean was at 144.7 BAU/mL and the levels dropped to 52.1 BAU/mL at T2. At one month after the MVC-COV1901 booster dose (T3), anti-spike IgG levels increased to 724.9 BAU/mL or an almost 14-fold increase compared to on the day of booster dose (T2). Compared to T1, boosting by a third dose of MVC-COV1901 elicited a 5.0-fold increase in anti-spike IgG immune response. As shown in Table 3, the neutralizing antibody (NAb) titer was increased from 59.0 IU/mL at the time of booster dose to 385.4 IU/mL at one month after the MVC-COV1901 booster dose. This amounted to 6.5-fold increase of NAb levels at T3 compared to T2 in terms of IU/mL or a 8.6-fold increase in NT_50_ (Table 3).

**Table 2.** Solicited Adverse Effects 14 days after an MVC-COV1901 booster shot

|  | Event | Participant<br>n (%) |
| --- | --- | --- |
| N (missing) |  | 100 (0) |
| At least one event | 257 | 82 |
| Local | 120 | 74(74.0) |
| Pain/Tenderness | 72 | 72(72.0) |
| Grade 1 | 71 | 71(71.0) |
| Grade 2 | 1 | 1(1.0) |
| Grade 3 | 0 | 0(0) |
| Erythema/Redness | 2 | 2(2.0) |
| Grade 1 | 2 | 2(2.0) |
| Grade 2 | 0 | 0(0) |
| Grade 3 | 0 | 0(0) |
| Induration/Swelling | 46 | 46(46.0) |
| Grade 1 | 46 | 46(46.0) |
| Grade 2 | 0 | 0(0) |
| Grade 3 | 0 | 0(0) |
| Systemic | 137 | 63(63.0) |
| Malaise/Fatigue | 51 | 51(51.0) |
| Grade 1 | 48 | 48(48.0) |
| Grade 2 | 3 | 3(3.0) |
| Grade 3 | 0 | 0(0) |
| Myalgia | 39 | 39(39.0) |
| Grade 1 | 35 | 35(35.0) |
| Grade 2 | 4 | 4(4.0) |
| Grade 3 | 0 | 0(0) |
| Headache | 30 | 30(30.0) |
| Grade 1 | 29 | 29(29.0) |
| Grade 2 | 1 | 1(1) |
| Grade 3 | 0 | 0(0) |
| Diarrhea | 7 | 7(7.0) |
| Grade 1 | 7 | 7(7.0) |
| Grade 2 | 0 | 0(0) |
| Grade 3 | 0 | 0(0) |
| Nausea/Vomiting | 10 | 10(10.0) |
| Grade 1 | 10 | 10(10.0) |
| Grade 2 | 0 | 0(0) |
| Grade 3 | 0 | 0(0) |
| Fever | 0 | 0(0) |
| Grade 1 | 0 | 0(0) |
| Grade 2 | 0 | 0(0) |
| Grade 3 | 0 | 0(0) |

**Table 3.** Immunogenicity at T2 (day of MVC-COV1901 booster) and T3 (one month after MVC-COV1901 booster) as measured by anti-SARS-CoV-2 spike IgG titers and live virus neutralizing antibody titers. GMTs are shown as GMT with 95% confidence interval in the parentheses. Fold changes are calculated as geometric mean of the T3/T2 ratio of individual titer values with 95% confidence interval in the parentheses.

|  | Unit | T2 | T3 | Fold change T3/T2 |
| --- | --- | --- | --- | --- |
| Anti-SARS-CoV-2 spike IgG | IgG GMT | 571.4 (456.6-715.1) | 7948.7 (6558.5-9633.6) | 13.9 (10.5-18.4) |
|  | BAU/mL GMT | 52.1 (41.6-65.2) | 724.9 (598.1-878.6) | 13.9 (10.5-18.4) |
| Neutralizing antibody | NT<sub>50</sub> GMT | 66.6 (56.6-78.4) | 569.7 (471.7-688.0) | 8.6 (7.0-10.5) |
|  | IU/mL GMT | 59.0 (51.2) | 385.4 (326.8-454.5) | 6.5 (5.5-7.8) |

Injection of a third shot of MVC-COV1901 was generally safe and no adverse effects greater than mild (grade 1) to moderate (grade 2) were reported. No serious adverse event was reported after a booster shot with MVC-COV1901 in participants who had received two immunizations with AZD1222. Table 2 presents the safety profile of the third dose of MVC-COV1901. Among recipients of the MVC-COV1901 booster shot, approximately 72% experienced pain or tenderness at the site of injection within 14 days after the shot. A noticeably lower proportion (i.e. 46%) of participants reported injection-site induration or swelling. Majority of those who encountered local solicited side effects, encountered grade 1 or mild effects while a small percentage experienced moderate reaction. In terms of systemic solicited AE, 51% experienced malaise or fatigue while none experienced fever. The most commonly reported systemic events were malaise or fatigue and myalgia (51% and 39%, respectively). Most systemic reactions reported were grade 1 with a few grade 2 reactions.

To test the neutralizing ability against the Omicron variant by antibodies induced by the MVC-COV1901 booster, we have randomly selected pre- and post-booster serum samples from 30 participants and subjected them to neutralizing assay against wildtype and Omicron variant pseudoviruses. Before the booster, two doses of AZD1222 were largely ineffective in neutralizing the Omicron variant pseudovirus at a 7.3-fold reduction in GMT compared to the wildtype, with only two individuals having detectable levels of neutralizing antibodies (Figure 4A). After the booster, about 90% of the individuals had detectable neutralizing titers against the Omicron variant pseudovirus. Compared to pre-booster levels, there were 5.2- and 5.7-fold increases in GMT levels against the wildtype and Omicron variant pseudoviruses compared to pre-booster levels, respectively (Figure 4B).

## Discussion

The antibody quantifications presented here demonstrate that following a waning period post second immunization with AZD1222, a booster shot with MVC-COV19 significantly raises binding antibody level by as much as 14-fold. More importantly, maximum antibody levels achieved with the booster exceed the maximum levels achieved by two immunizations with AZD1222. Similar results have been observed in an extension of the MVC-COV1901 phase 1 trial that followed the same observation and vaccination time points as this study, but used MVC-COV1901 for all three immunizations [13, 14].

The aggregated results from the phase 1 study and the study presented here, suggest that options of a booster vaccine may not be limited to the matching vaccines for the primary series. More importantly, boosters may have the general property of efficiently raising neutralizing antibody to levels not achieved by prior immunizations [15, 16]. Based on the correlates of protection published from Phase III data of AZD1222, the predicted VE against symptomatic infection against the ancestral strain at four months after the second AZD1222 dose was below 60%, after the booster the predicted VE was raised to 89.7%. [17]. There is, however, an important follow up question that is to determine if the neutralizing antibody levels are better maintained long term after one or more boosters. The recent surges of infections driven by VoCs such as Delta and Omicron variants were alarming in that two doses of currently available vaccines are largely ineffective against the Omicron variant in particular [4, 18]. As more data came to light on the effectiveness of booster doses to improve neutralizing against the Omicron variant and other VoCs, booster doses remain the only viable option at this point to mitigate the waves of infections [18-20]. In this study, we have shown that a booster dose of MVC-COV1901 after two doses of AZD1222 can result in similar increases in neutralizing titer levels against the wildtype and Omicron variant pseudoviruses (Figure 4). Administration of booster dose restored the neutralizing ability against the Omicron variant that was previously undetectable with only two doses of vaccination, as in line with other booster studies [18-20]. Recent real world results of three doses of mRNA-1273 against the VoCs showed that the vaccine efficacies against Delta and Omicron infection were 95.2% and 62.5%, respectively [18]. The immunogenic response and favorable safety profile of the third booster shot might offer a viable option for vaccination program designers to cope with the issue of waning immunity after the two-dose schedule compounded by the emergence of variants of concern. The outcomes of the MVC-COV1901 booster remains to be seen in the real world efficacy study.

Recent study in the UK for heterologous third dose booster regimen (COV-BOOST) showed that at one month after booster dose compared to the day of the booster dose, there was a 2.6-fold increase in anti-spike IgG titer with two doses plus a booster dose of AZD1222 and a 6.7-fold increase with two doses of AZD1222 and a booster dose of Novavax NVX-CoV2373, also a protein subunit vaccine [21]. In our study, a booster dose of MVC-COV1901 after two doses of AZD1222 showed a 14-fold increase in anti-spike IgG titer, superior to that of the above two combinations and nearly equal to that of boosting with BNT162b2 after two doses of AZD1222, which achieved a 16-fold increase in the COV-BOOST study [21]. In terms of reactogenicity, we have observed no occurrence of severe (grade 3) adverse events after booster administration, while severe solicited reactions have been reported for all of heterologous booster combinations in the COV-BOOST study [21]. Most noticeably, no incidence of fever has been reported in this study, similar to our previous clinical findings [11].

From our interim findings, we showed that administration of MVC-COV1901 as a third dose booster could effectively and safely enhance immunogenicity nearly to the level as seen in using mRNA vaccine as booster dose. Our strategy could be aptly applied to lower and middle income countries that would have difficulties in handling mRNA booster vaccines but desirable to achieve a similarly high level of immunity against SARS-CoV-2.

## Data Availability

All data produced in the present study are available upon reasonable request to the authors

## Author Contributions

Concept and design: S.-H.C.

Acquisition and interpretation/analysis of data, manuscript preparation: S.-H.C., Y.-C.L., C.-P.C., and C.-Y. C.

## Additional Contributions

The Institute of Biomedical Sciences, Academia Sinica performed the neutralization assay. Members of Medigen Vaccine Biologics Corp. assisted in manuscript editing and revision.

## Ethics Statement

The trial protocol and informed consent form were approved by the Medical Ethics and Institutional Review Board of Taoyuan General Hospital. The trial was conducted in accordance with the principles of the Declaration of Helsinki and Good Clinical Practice. This trial is registered on ClinicalTrials.gov: NCT05097053.

